# Clarifying predictions for COVID-19 from testing data: the example of New-York State

**DOI:** 10.1101/2020.10.10.20203034

**Authors:** Quentin Griette, Pierre Magal

**Affiliations:** Univ. Bordeaux, IMB, UMR 5251, F-33400 Talence, France; CNRS, IMB, UMR 5251, F-33400 Talence, France

**Author notes:** ANR flash COVID-19: MPCUII.

**Keywords:** corona virus, testing data, reported and unreported cases, isolation, quarantine, public closings, epidemic mathematical model

## Abstract

In this article, we use testing data as an input of a new epidemic model. We get nice a concordance between the best fit the model to the reported cases data for New-York state. We also get a good concordance of the testing dynamic and the epidemic’s dynamic in the cumulative cases. Finally, we can investigate the effect of multiplying the number of tests by 2, 5, 10, and 100 to investigate the consequences on the reduction of the number of reported cases.

## 1 Introduction

The epidemic of novel coronavirus (COVID-19) infections began in China in December 2019 and rapidly spread worldwide in 2020. Since the early beginning of the epidemic, mathematicians and epidemiologists have developed models to analyze the data and characterize the spread of the virus, and attempt to project the future evolution of the epidemic. Many of those models are based on the SIR or SEIR model which is classical in the context of epidemics. We refer to [26, 28] for the earliest article devoted to such a question and we refer to [1, 3–7, 10, 12, 13, 20, 25] for a rather complete overview on SIR and SEIR models in general. In the course of the COVID-19 outbreak, it became clear for the scientific community that covert cases (asymptomatic or unreported infectious case) play an important role. An early description of an asymptomatic transmission in Germany was reported by Rothe et al. [24]. It was also observed on the Diamond Princess cruise ship in Yokohama in Japan by Mizumoto et al. [19] that many of the passengers were tested positive to the virus, but never presented any symptoms. We also refer to Qiu [21] for more information about this problem. At the early stage of the COVID-19 outbreak, a new class of epidemic models was proposed in Liu et al. [14] to take into account the contamination of susceptible individuals by contact with unreported infectious. Actually, this class of model was presented earlier in Arino et al. [2]. In [14] a new method to use the number of reported in SIR models was also proposed. This method and model was extended in several directions by the same group in [15–17] to include non-constant transmission rates and a period of exposure. More recently the method was extended and successfully applied to a Japanese age-structured dataset in [11]. The method was also extended to investigate the predictability of the outbreak in several countries China, South Korea, Italy, France, Germany and the United Kingdom in [18]. The application of the Bayesian method was also considered in [9].

In parallel with these modeling ideas, Bayesian methods have been widely used to identify the parameters in the models used for the COVID-19 pandemic (see e.g. Roques et al. [22,23] where an estimate of the fatality ratio has been developed). A remarkable feature of those methods is to provide mechanisms to correct some of the known biases in the observation of cases, such as the daily number of tests. Here we will embed the data for the daily number of tests into an epidemic model, and we will compare the number of reported cases produced by the model and the data. Our goal is to understand the relationship between the data for the daily number of tests (which will be an input our model) and the data for the daily number of reported cases (which will be an output for our model).

The plan of the paper is the following. In Section 2, we will present a model involving the daily number of tests. In Section 3, we apply the method presented in [14] to our new model. In Section 4, we present some numerical simulations, and we compare the model with the data. The last section is devoted to the discussion.

## 2 Epidemic with testing data

Let *n*(*t*) be the number of tests per unit of time. Throughout this paper, we use one day as the unit of time. Therefore *n*(*t*) can be regarded as the daily number of tests at time *t*. The function *n*(*t*) is actually coming from a database for the New-York State [29]. Let *N* (*t*) be the cumulative number of tests from the beginning of the epidemic then

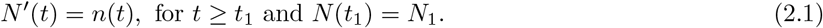

### Remark 2.1

*Section 4 is devoted numerical simulations. We will use n*(*t*) *as a piecewise constant function that varies day by day. Each day, n*(*t*) *will be equal to the number of tests that were performed that day. So n*(*t*) *should be understood as the black curve in Figure 4*.

**Figure 1:**
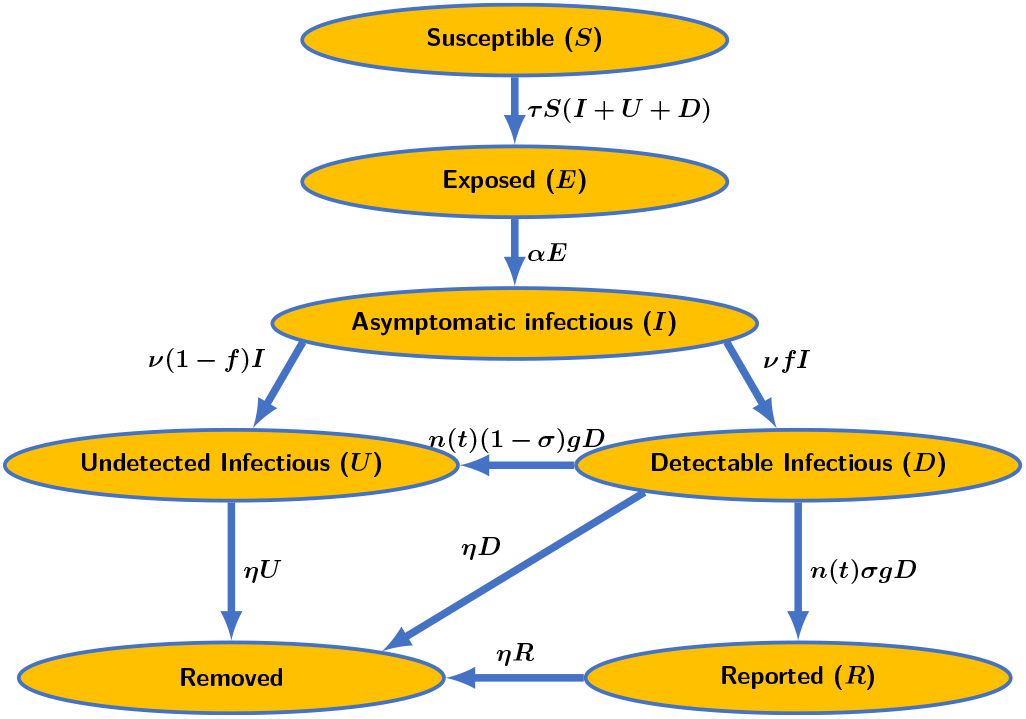
Flow chart of the epidemic model with tests (2.2). In this diagram n(t) is the daily number of tests at time t. We consider a fraction (1 − σ) of false negative tests and a fraction σ of true positive tests. The parameter g reflects the fact that the tests are not only devoted only to the symptomatic patients but to a large fraction of the population of New-York state.

**Figure 2:**
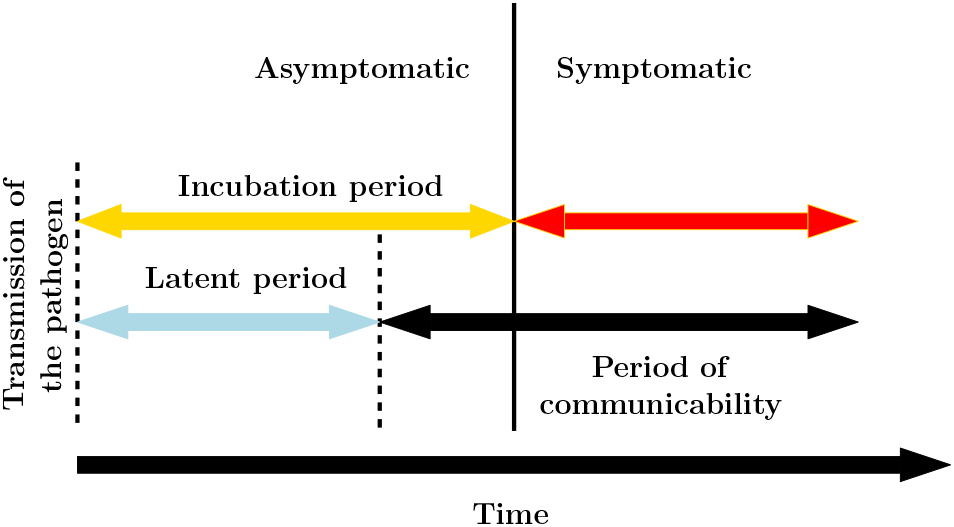
Key time periods of COVID-19 infection: the latent or exposed period before the onset of symptoms and transmissibility, the incubation period before symptoms appear, the symptomatic period, and the transmissibility period, which may overlap the asymptomatic period.

**Figure 3:**
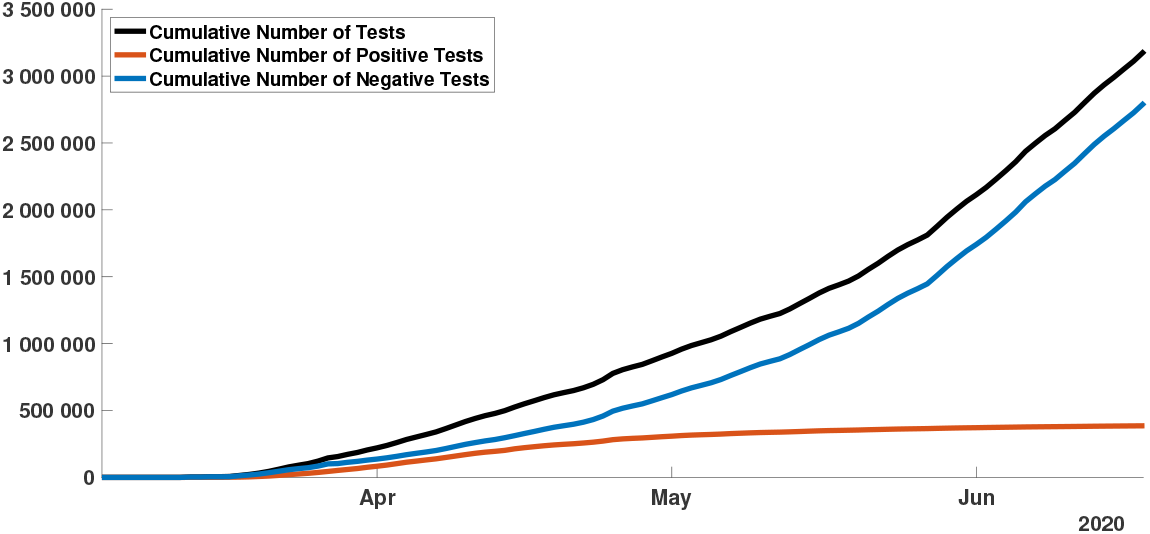
In this figure, we plot the cumulative number of tests for the New-York State. The black curve, orange curve, and blue curve correspond respectively to the number of tests, the number of positive tests, and the number of negative tests. We can see that at the early beginning of the epidemic, the cumulative number of tests (black curve) grows linearly approximatively from mid-March to mid-April.

**Figure 4:**
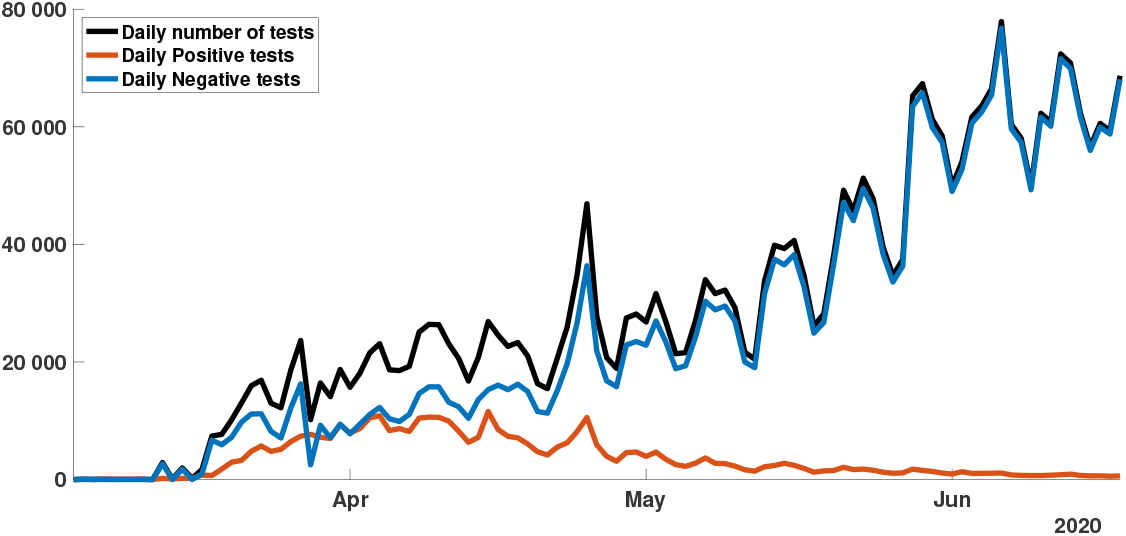
In this figure, we plot the daily number of tests for the New-York State. The black curve, orange curve, and blue curve correspond respectively to the number of tests, the number of positive tests, and the number of negative tests.

The model consists of the following system of ordinary differential equations

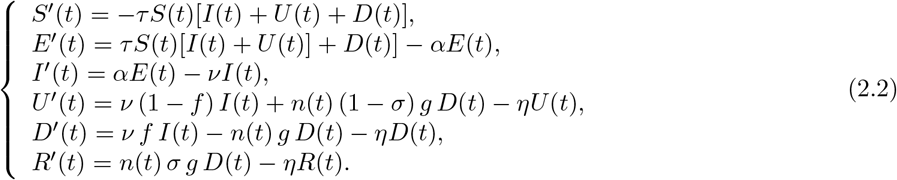

This system is supplemented by initial data (which are all non negative)

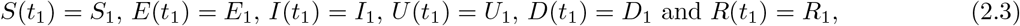

thereby assuming that the disease was introduced by an individual incubating the disease at some time before *t*^_1_^. The time *t*_1_ corresponds to the time where the tests started to be used constantly. Therefore the epidemic started before *t*_1_.

Here *t* ≥ *t*_1_ is the time in days. *S*(*t*) is the number of individuals susceptible to infection. *E*(*t*) is the number of exposed individuals (*i*.*e*. who are incubating the disease but not infectious). *I*(*t*) is the number of individuals incubating the disease, but already infectious. *U* (*t*) is the number of undetected infectious individuals (*i*.*e*. who are expressing mild or no symptoms), and the infectious that have been tested with a false negative result, are therefore not candidates for testing. *D*(*t*) is the number of individuals who express severe symptoms and are candidates for testing. *R*(*t*) is the number of individuals who have been tested positive to the disease. The flux diagram of our model is presented in Figure 1.

Susceptible individuals *S*(*t*) become infected by contact with an infectious individual *I*(*t*), *U* (*t*) or *D*(*t*). When they get infected, susceptible are first classified as exposed individuals *E*(*t*), that is to say that they are incubating the disease but not yet infectious. The average length of this exposed period (or noninfectious incubation period) is 1*/α* days.

After the exposure period, individuals are becoming asymptomatic infectious *I*(*t*). The average length of the asymptomatic infectious period is 1*/ν* days. After this period, individuals are becoming either mildly symptomatic individuals *U* (*t*) or individuals with severe symptoms *D*(*t*). The average length of this infectious period is 1*/η* days. Some of the *U* -individuals may show no symptoms at all.

In our model, the transmission can occur between a *S*-individual and an *I*-, *U* - or *R*-individual. Transmissions of SARS-CoV-2 are described in the model by the term *τ S*(*t*)[*I*(*t*) + *U* (*t*) + *D*(*t*)] where *τ* is the transmission rate. Here, even though a transmission from *R*-individuals to a *S*-individuals is possible in theory (e.g. if a tested patient infects its medical doctor), we consider that such a case is rare and we neglect it.

The last part of the model is devoted to the testing. The parameter *σ* is the fraction of true positive tests and (1 − *σ*) is the fraction of false negative tests. The quantity *σ* has been estimated at *σ* = 0.7 in the case of nasal or pharyngeal swabs for SARS-CoV-2 [27].

Among the detectable infectious, we assume that only a fraction *g* are tested per unit of time. This fraction corresponds to individuals with symptoms suggesting a potential infection to SARS-CoV-2. The fraction *g* is the frequency of testable individuals in the population of New-York state. We can rewrite *g* as

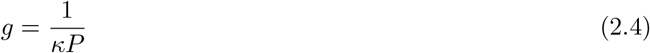

where *P* is the total number of individuals in the population of the state of New-York and 0 ≤ *κ* ≤ 1 is the fraction total population with mild or sever symptoms that may induce a test.

Individuals who were tested positive *R*(*t*) are infectious on average during a period of 1*/η* days. But we assume that they become immediately isolated and they do not contribute to the epidemic anymore. In this model we focus on the testing of the *D*-individuals. The quantity *n*(*t*) *σ g D* is flux of successfully tested *D*-individuals which become *R*-individuals. The flux of tested *D*-individuals which are false negatives is *n*(*t*) (1 − *σ*) *g D* which go from the class of *D*-individuals to the *U* -individuals. The parameters of the model and the initial conditions of the model are listed in Table 1.

**Table 1:** Parameters and initial conditions of the model.

| Symbol | Interpretation | Method |
| --- | --- | --- |
| $t_1$ | Date when the tests start to be used extensively | fixed |
| $S_1$ | Number of susceptible at time $t_1$ | fixed |
| $E_1$ | Number of exposed at time $t_1$ | fitted |
| $I_1$ | Number of asymptomatic infectious at time $t_1$ | fitted |
| $U_1$ | Number of undetectable infectious at time $t_1$ | fitted |
| $D_1$ | Number of detectable infectious at time $t_1$ | fitted |
| $R_1$ | Number of reported (tested positive) cases at time $t_1$ | fitted |
| $\tau$ | Transmission rate | fitted |
| $n(t)$ | Number of tests per unit of time | fixed |
| $1/\alpha$ | Average length of exposure | fixed |
| $1/\nu$ | Average length of asymptomatic infectiousness | fixed |
| $1/\eta$ | Average length of symptomatic infectiousness | fixed |
| $f$ | Frequency of infectious with severe symptoms | fixed |
| $\sigma$ | Fraction of true positive tests | fixed |
| $g$ | Frequency of testable individuals | fixed |

**Table 2:**
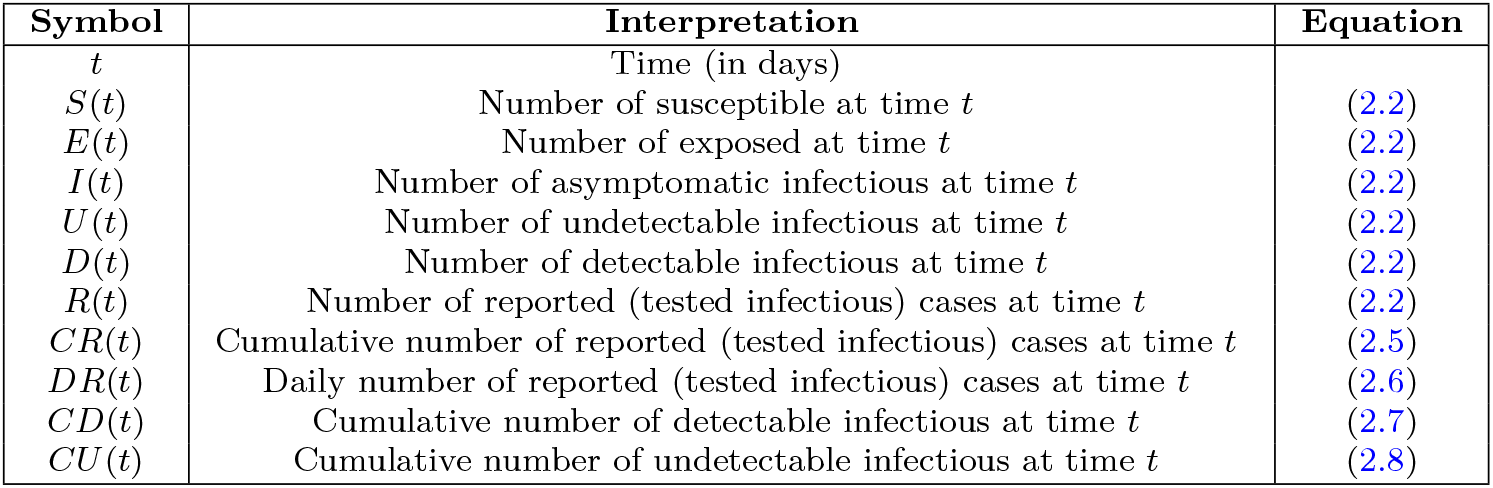
Variables used in the models.

| Symbol | Interpretation | Equation |
| --- | --- | --- |
| $t$ | Time (in days) | |
| $S(t)$ | Number of susceptible at time $t$ | (2.2) |
| $E(t)$ | Number of exposed at time $t$ | (2.2) |
| $I(t)$ | Number of asymptomatic infectious at time $t$ | (2.2) |
| $U(t)$ | Number of undetectable infectious at time $t$ | (2.2) |
| $D(t)$ | Number of detectable infectious at time $t$ | (2.2) |
| $R(t)$ | Number of reported (tested infectious) cases at time $t$ | (2.2) |
| $CR(t)$ | Cumulative number of reported (tested infectious) cases at time $t$ | (2.5) |
| $DR(t)$ | Daily number of reported (tested infectious) cases at time $t$ | (2.6) |
| $CD(t)$ | Cumulative number of detectable infectious at time $t$ | (2.7) |
| $CU(t)$ | Cumulative number of undetectable infectious at time $t$ | (2.8) |

Before describing our method we need to introduce a few useful identities. The cumulative number of reported cases is obtained by using the following equation

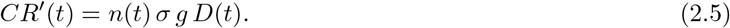

The daily number of reported cases *DR*′(*t*) is given by

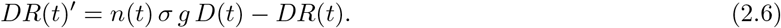

The cumulative number of detectable cases is given by

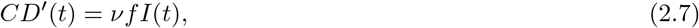

and the cumulative number of undetectable cases is given by

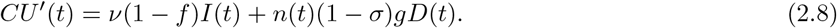

## 3 Method to fit the cumulative number of reported cases

In order to deal with data, we need to understand how to set the parameters as well as some components of the initial conditions. In order to do so, we extend the method presented first in [14]. The main novelty here will concern the cumulative number of tests which is assumed to grow linearly at the beginning. This property is satisfied for the New-York State data as we can see in Figure 3. The black curve in this figure is close to a line from March 15 to April 15. Figure 4 shows day-by-day fluctuations of the number of tests while in Figure 3 the day-by-day fluctuations are not visible and the cumulative data allow to understand the growth tendency of the number of tests.

### Phenomenological models for the tests

We fit a line to the cumulative number of tests in a suitable interval of days [*t*_1_, *t*_2_]. This means that we can find a pair of numbers *a* and *b* such that

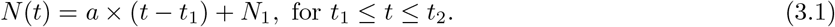

where *a* the daily number of tests and *N*_1_ is the cumulative number of tests on day *t*_1_.

By using the fact that *N* (*t*)′ = *n*(*t*) we deduce that

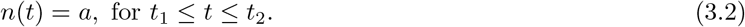

#### Remark 3.1

*In the simulations we will fit a line to the cumulative number of tests from mid-March to mid-April. Figure 3 shows that the linear growth assumption is reasonable for the New-York State cumulative testing data*.

### Phenomenological models for the reported cases

At the early stage of the epidemic, we assume that all the infected components of the system grow exponentially while the number of susceptible remains unchanged during a relatively short period of time *t* ∈ [*t*_1_, *t*_2_]. Therefore, we will assume that

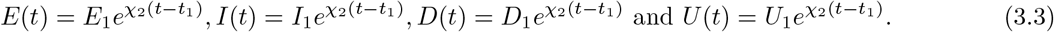

We deduce that the cumulative number of reported satisfies

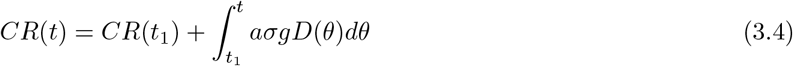

hence by replacing *D*(*t*) by the exponential formula (3.3)

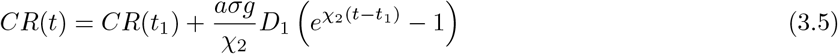

and it is makes sense to assume that *CR*(*t*) − *CR*(*t*_1_) has the following form

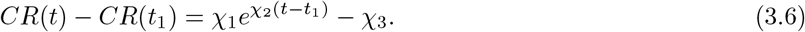

By identifying (3.5) and (3.6) we deduce that

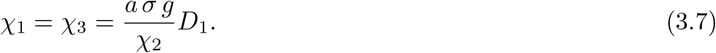

Moreover by using (3.2) and the fact that the number of susceptible *S*(*t*) remains constant equal to *S*_1_ on the time interval *t* ∈ [*t*_1_, *t*_2_], the *E*-equation, *I*-equation, *U* -equation and *D*-equation of the model (2.2) become

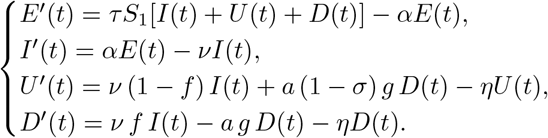

By using (3.3) we obtain

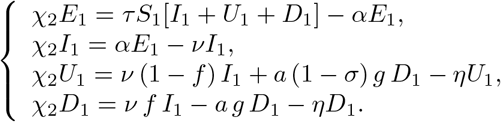

Computing further, we get

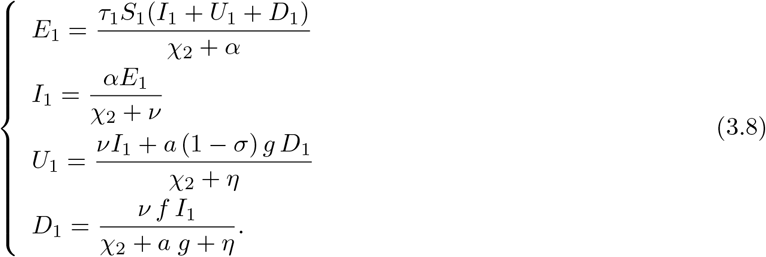

Finally by using (3.7)

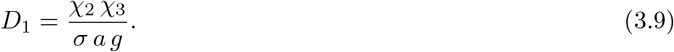

and by using (3.8) we obtain

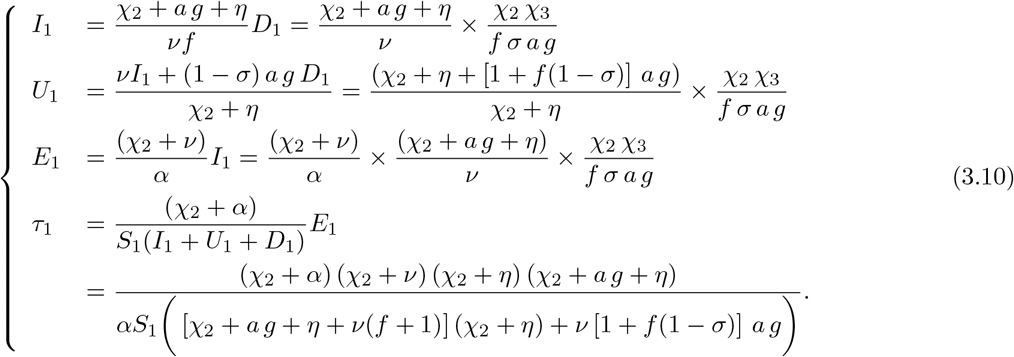

## 4 Numerical simulations

We assume that the transmission coefficient takes the form

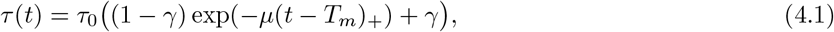

where *τ*_0_ > 0 is the initial transmission coefficient, *T*_*m*_ > 0 is the time at which the social distancing starts in the population, *µ* > 0 is serving to modulate the speed at which this social distancing is taking place.

To take into account the effect of social distancing and public measures, we assume that the transmission coefficient *τ* (*t*) can be modulated by *γ*. Indeed by the closing of schools and non-essential shops and by imposing social distancing the population of the New-York State, the number of contacts per day is reduced. This effect was visible on the news during the first wave of the COVID-19 epidemic in New-York city since the streets were almost empty at some point. The parameter *γ* > 0 is the percentage of the number of transmissions that remain after a transition period (depending on *µ*), compared to a normal situation. A similar non-constant transmission rate was considered by Chowell et al. [8].

In Figure 5 we consider a constant transmission rate *τ* (*t*) ≡ *τ*_0_ which corresponds to *γ* = 1 in (4.1). In order to evaluate the distance between the model and the data, we compare the distance between the cumulative number of cases *CR* produced by the model and the data (see the orange dots and orange curve in Figure 5-(a)). In Figure 5-(c) we can observe that the cumulative number of cases increases up more than 14 millions of people, which indeed is not realistic. Nevertheless by choosing the parameter 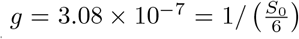 in Figure 5-(d) we can see that the orange dots and the blue curve match very well.

**Figure 5:**
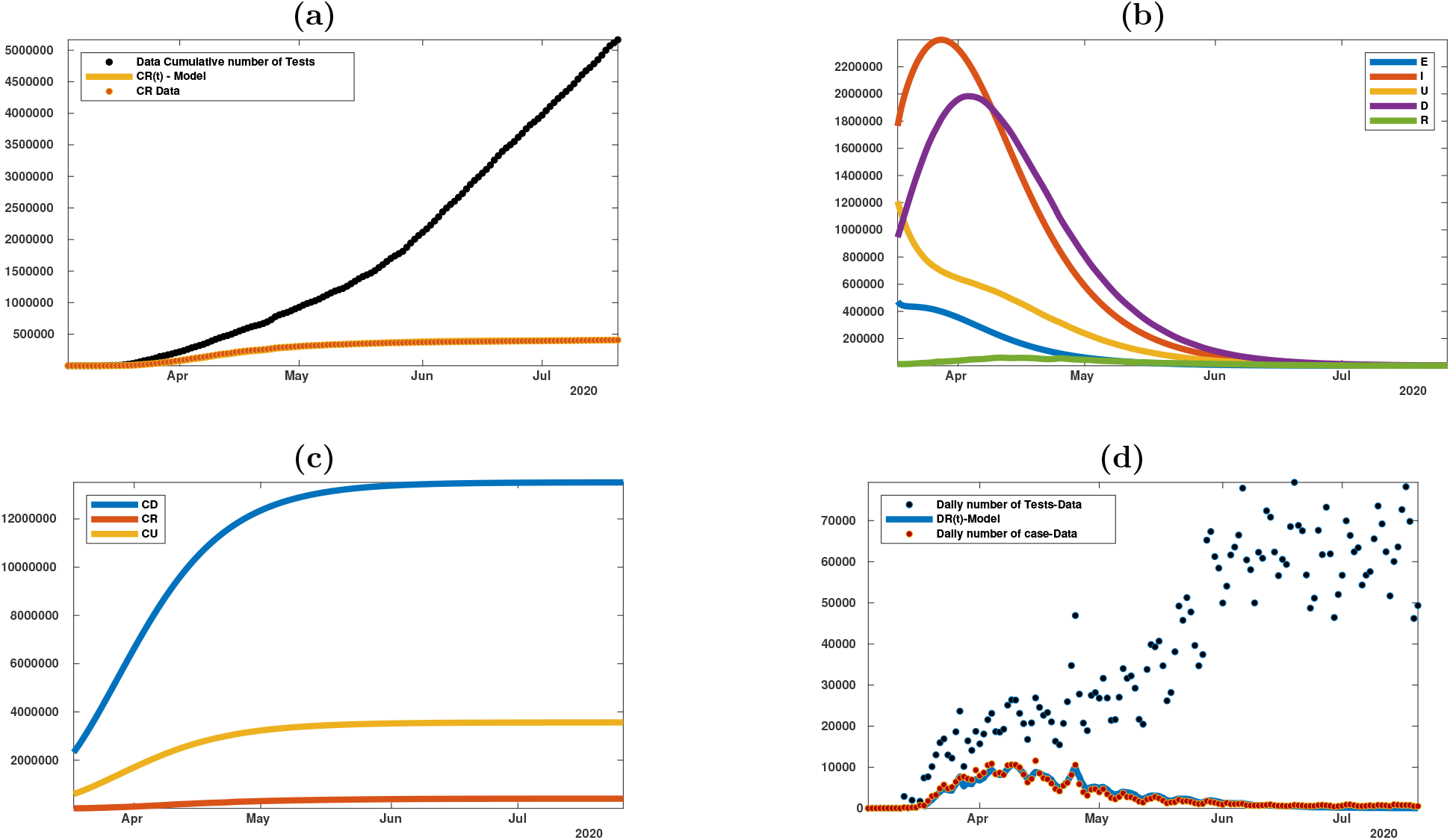
Best fit of the model without confinement (or social distancing) measures (i.e. γ = 1). **Fitted parameters:** The transmission rate τ (t) ≡ τ_0_ is constant according to the formula (4.1) whenever γ = 1 and τ_0_ is fitted by using (3.10). **Parameter values:** S_0_ = 19453561, α = 1, ν = 1/6, η = 1/7, σ = 0.7, f = 0.8 and g = 6/S_0_ = 3.08 × 10^−7^. t_1_ = march 18, t_2_ = march 29, a = 1.4874 × 10^4^, b = − 2.1781 × 10^5^, χ_1_ = 2.8814 × 10^4^, χ_2_ = 0.1013, χ_3_ = 2.9969 × 10^4^. The parameter τ_0_ is fitted by using (3.10). In figure **(a)** we plot the cumulative number of tests (black dots), the cumulative number of positive cases (red dots) for the state of New-York and the cumulative number of cases CD(t) (yellow curve) obtained by fitting the model to the data. In figures **(b)–(c)** we plot the number of cases obtained from the model. We can observe that most of the cases are unreported. In figure **(d)** we plot the daily number of tests (black dots), the daily number of positive cases (red dots) for the state of New-York and the daily number of cases DD(t) obtained from the data.

In the rest of this section, we focus on the model with confinement (or social distancing) measures. We assume that such social distancing measures have a strong impact on the transmission rate by assuming that *γ* = 0.2 < 1. It means that only 20% of the transmissions will remain after a transition period.

In Figure 6-(c) we can observe that the cumulative number of cases increases up to 800 000 (blue curve) while the cumulative number reported cases goes up to 350 000. In Figure 6-(d) we can see that the orange dots and the blue curve match very well again. In order to get this fit we fix the parameter *g* = 10^−5^.

**Figure 6:**
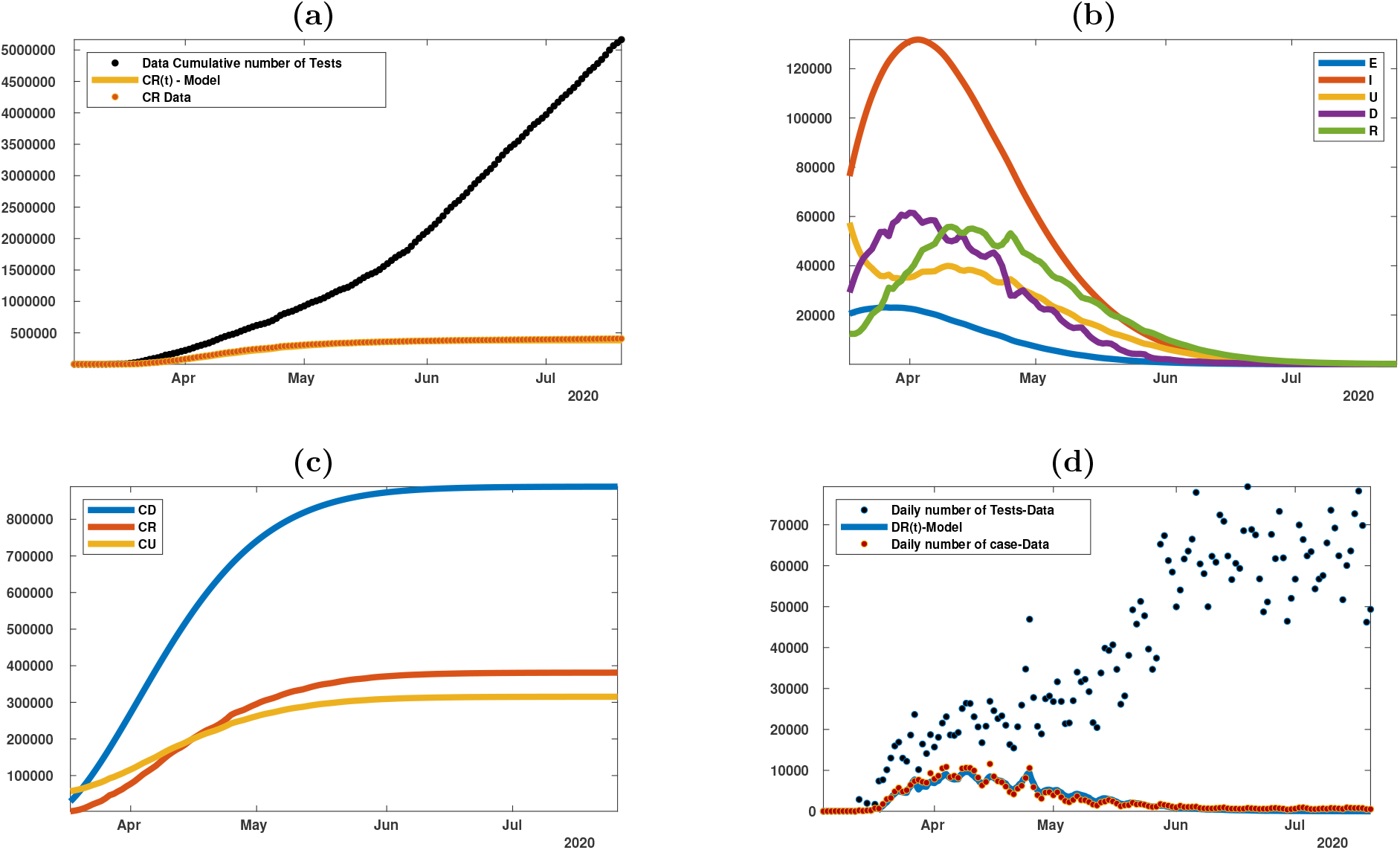
Best fit of the model with confinement (or social distancing) measures. **Parameter values:** Same in Figure 5, excepted the transmission coefficient which is not constant in time with γ = 0.2 and τ_0_ is fitted by using (3.10), T_m_ = 15 Mar (starting day of public measures), µ = 0.0251 and τ_0_ is fitted by using (3.10) and g = 10^−5^. In figure **(a)** we plot the cumulative number of tests (black dots), the cumulative number of positive cases (red dots) for the state of New-York and the cumulative number of cases CD(t) (yellow curve) obtained by fitting the model to the data. In figures **(b)–(c)** we plot the corresponds number of cases obtained from the model. With this set of parameters we can check that most of the cases are unreported. In figure **(d)** we plot the daily number of tests (black dots), the daily number of positive cases (red dots) for the state of New-York and the daily number of cases DD(t) obtained from the data.

In Figure 7 (a) and (b), we aim at understanding the connection between the daily fluctuations of the number of reported cases (epidemic dynamic) and the daily number of tests (testing dynamics). The combination of both the testing dynamics and the infection dynamics gives indeed a very complex curve parametrized by the time. It seems that the only reasonable comparison that we can make is between the cumulative number of reported cases and the cumulative number of tests. In Figure 7 (c) and (d), the comparison of the model and the data gives a very decent fit.

**Figure 7:**
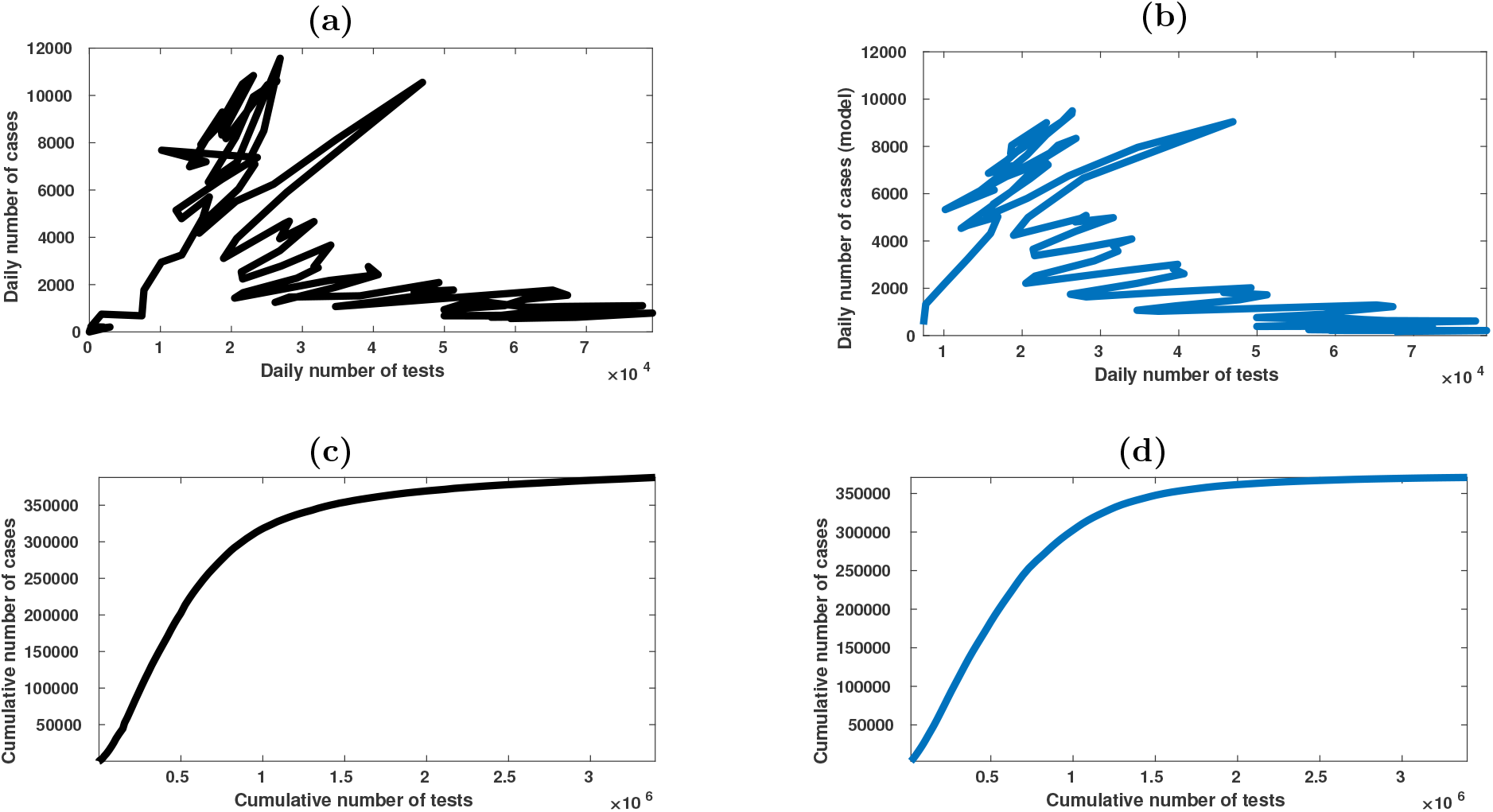
In this figure we plot the curves of the the number of reported cases in function of the number of tests parametrized by the time. The top figures (a) and (b) correspond to the daily number of cases and the bottom figures (c) and (d) correspond to the cumulative number of cases. On the left-hand side we plot the data (a) and (c) while on the right-hand side we plot the model (b) and (d). **Parameter values:** are the same as in Figure 6. In figure **(a)** we plot the daily number of cases coming from the data as a function of the daily number of tests. In figure **(b)** we plot the daily number of cases given by the model as a function of the cumulative number of cases coming from the data. In figure **(c)** we plot the cumulative number of cases coming from the data as a function of the cumulative number of tests. In figure **(d)** we plot the cumulative number of cases coming from the model as a function of the cumulative number of tests from the data.

In Figure 7, all the curves are time dependent parametrized curves. The abscissa is the number of tests (horizontal axis) and the ordinate is the number of reported cases (vertical axis). It corresponds (with our notations) to the parametric functions *t* → (*n*_*data*_(*t*), *DR*(*t*)) in figures (a) and (b) and their cumulative equivalent *t* → (*N*_*data*_(*t*), *CR*(*t*)) in figure (c) and (d). In figures (a) and (c) we use only the data, that is to say that we plot *t* → (*n*_*data*_(*t*), *DR*_*data*_(*t*)) and *t* → (*N*_*data*_(*t*), *CR*_*data*_(*t*)). In figures (b) and (d) we use only the model for the number of reported cases, that is to say that we plot *t* → (*n*_*data*_(*t*), *DR*_*model*_(*t*)) and *t* → (*N*_*data*_(*t*), *CR*_*model*_(*t*)).

In Figure 8, our goal is to investigate the effect of a change in the testing policy in the New-York State. We are particularly interested in estimating the effect of an increase of the number of tests on the epidemic. Indeed people commonly say that increasing the number of tests will be beneficial to reduce the number of cases. So here, we try to quantify this idea by using our model.

**Figure 8:**
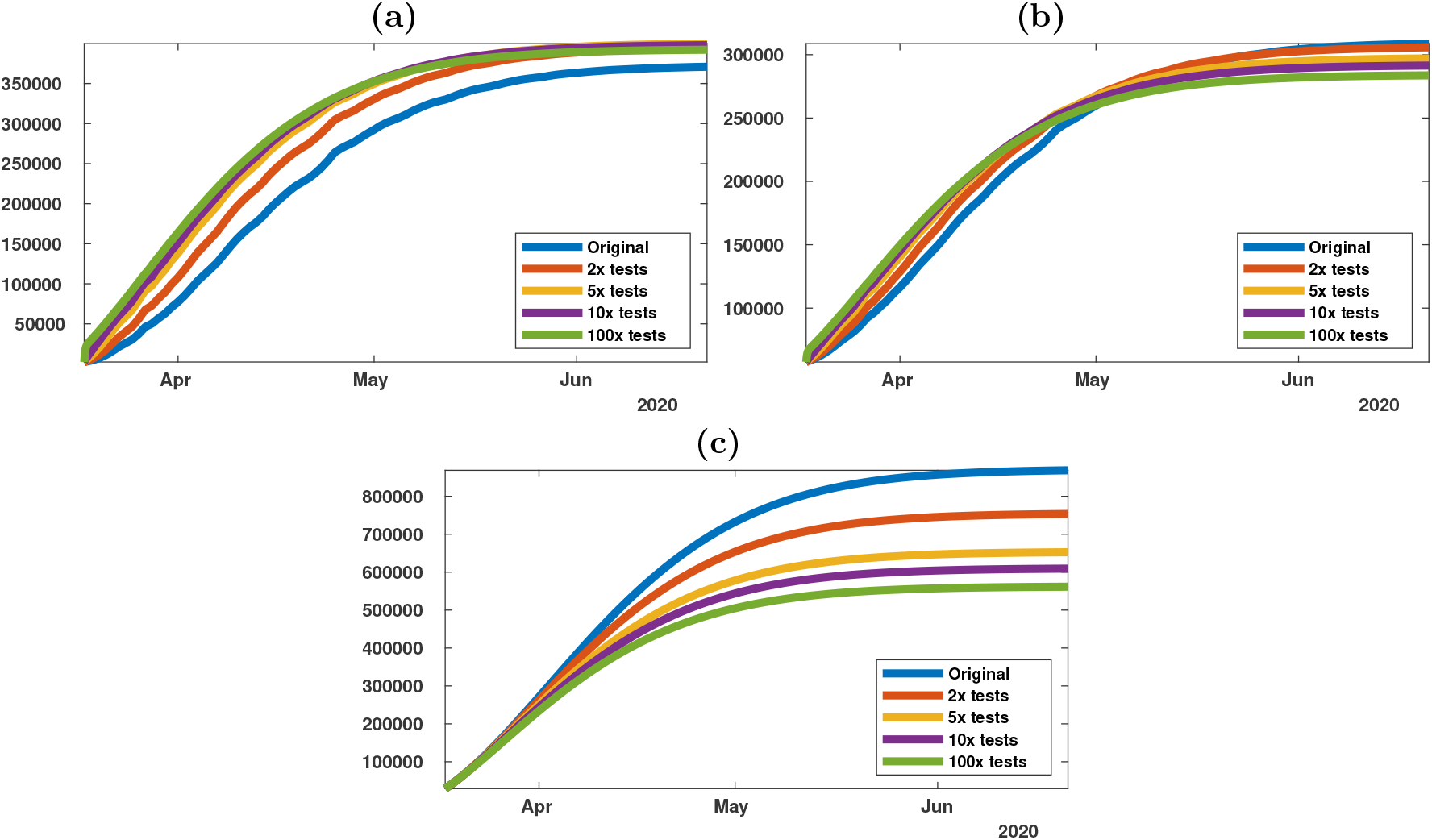
Cumulative number of cases for different testing strategies: Original (blue curve), doubled (red curve), multiplied by 5 (yellow curve), multiplied by 10 (purple line) and multiplied by 100 (green curve). The transmission coefficient depends on the time, according to the formula (4.1) with γ = 0.2 and τ_0_ is fitted by using (3.10). **Parameter values:** are the same as in Figure 6. In figure **(a)** we plot the cumulated number of cases CR(t) as a function of time. In figure **(b)** we plot the cumulative number of undetectable cases CU (t) as a function of time. In figure **(c)** we plot the cumulative number of cases (including covert cases) CD(t) as a function of time. Note that the total number of cases (including covert cases) is reduced by 35% when the number of tests is multiplied by 100.

In Figure 8, we replace the daily number of tests *n*_*data*_(*t*) (coming from the data for New-York’s state) in the model by either 2 × *n*_*data*_(*t*), 5 × *n*_*data*_(*t*), 10 × *n*_*data*_(*t*) or 100 × *n*_*data*_(*t*).

As expected, an increase of the number of tests is helping to reduce the number of cases. However, after increasing 10 times the number of tests, there is no significant difference (in the number of reported) between 10 times and 100 times more tests. Therefore there must be an optimum between increasing the number of tests (which costs money and other limited resources) and being efficient to slow down the epidemic.

## 5 Discussion

In this article, we propose a new epidemic model involving the daily number of tests as an input of the model. The model itself is extending our previous models presented in [11, 14–18]. We propose a new method to use the data in such a context based on the fact that the cumulative number of tests grows linearly at the early stage of the epidemic. Figure 3 shows that this is a reasonable assumption for the New-York State data from mid-March to mid-April.

Our numerical simulations show a very good concordance between the number of reported cases produced by the model and the data in two very different situations. Indeed, Figures 5 and 6 correspond respectively to an epidemic without and with public intervention to limit the number of transmissions. This is an important observation since this shows that testing data and reported cases are not sufficient to evaluate the real amplitude of the epidemic. To solve this problem, the only solution seems to include a different kind of data to the models. This could be done by studying statistically representative samples in the population. Otherwise, biases can always be suspected. Such a question is of particular interest in order to evaluate the fraction of the population that has been infected by the virus and their possible immunity.

In Figure 7, we compare the testing dynamic (day by day variation in the number of tests) and the reported cases dynamic (day by day variation in the number of reported). Indeed, the daily case is extremely complex, but we also obtain some relatively robust curve for the cumulative numbers. Our model give a good fit for this cumulative cases.

In Figure 8, we compare multiple testing strategies. By increasing 2, 5, 10 and 100 times the number of tests, we can observe that this efficient up to some point 10 and but increasing 100 times is not making a big difference. Therefore, it is useless to test to many peoples and there must an optimum (between the cost of the tests) and the efficiency in the evaluation of the number of cases.

## Data Availability

The data used are public. See the references in the paper

## Acknowledgements

Data from [29].

